# Automating intended target identification for paraphasias in discourse using a large language model

**DOI:** 10.1101/2023.06.18.23291555

**Authors:** Alexandra C. Salem, Robert C. Gale, Mikala Fleegle, Gerasimos Fergadiotis, Steven Bedrick

## Abstract

**Purpose:** To date there are no automated tools for the identification and fine-grained classification of paraphasias within discourse, the production of which is the hallmark characteristic of most people with aphasia (PWA). In this work we fine-tune a large language model (LLM) to automatically predict paraphasia targets in Cinderella story retellings.

**Method:** Data consisted of 353 Cinderella story retellings containing 2,489 paraphasias from PWA, for which research assistants identified their intended targets. We supplemented this training data with 256 sessions from control participants, to which we added 2,427 synthetic paraphasias. We conducted four experiments using different training data configurations to fine-tune the LLM to automatically “fill in the blank” of the paraphasia with a predicted target, given the context of the rest of the story retelling. We tested the experiments’ predictions against our human-identified targets and stratified our results by ambiguity of the targets and clinical factors.

**Results:** The model trained on controls and PWA achieved 46.8% accuracy at exactly matching the human-identified target. Fine-tuning on PWA data, with or without controls, led to comparable performance. The model performed better on targets with less human ambiguity, and on paraphasias from participants with less severe or fluent aphasia.

**Conclusion:** We were able to automatically identify the intended target of paraphasias in discourse using just the surrounding language about half of the time. These findings take us a step closer to automatic aphasic discourse analysis. In future work, we will incorporate phonological information from the paraphasia to further improve predictive utility.

---

Anomia or word-finding difficulty is a prominent and persistent feature of aphasia (Goodglass and Wingfield, 1997) and manifests in all communicative contexts, from single word responses to complex conversations. Given the ubiquitous nature of anomia, anomia assessments are given in most clinical settings and are of high practical value for quantifying performance and monitoring outcomes. Typically, anomia assessments include confrontation picture naming tests (Rabin et al., 2005; Simmons-Mackie, Threats, & Kagan, 2005), in which a person with aphasia is asked to name a series of pictured objects and/or actions. The popularity of confrontation picture naming tests can be attributed to their well-documented validity and reliability (e.g., Roach et al., 1996; Strauss, Sherman, & Spreen, 2006; Walker & Schwartz, 2012), and also to their relatively low testing burden, particularly in the context of short forms and simple accuracy scoring schemes. Other sources of diagnostic information such as discourse-level analyses may provide additional clinically useful information for completing a patient’s clinical profile (Fergadiotis et al., 2019; Richardson et al., 2018) but such analyses are not performed routinely in clinical settings. Viewed through an implementation science lens (Damschroder et al., 2009; Breimaier et al., 2015), several barriers hinder the utilization of discourse-based analyses including their complexity, reliability, and time burden. The latter factor especially can be an insurmountable barrier for implementation in most real-world clinical settings. Therefore, there is a need to develop new approaches that will enable professionals to assess people with aphasia (PWA) in a more objective, precise, efficient, and ecologically valid manner.

Computational methods, especially those from the field of Natural Language Processing (NLP), have the potential to be essential tools in designing such approaches. Recent work has demonstrated these methods’ efficacy in automating certain aspects of confrontation naming test scoring (Casilio et al., 2023; Salem et al., 2022; Fergadiotis et al., 2016; McKinney-Bock & Bedrick, 2019; described later in more detail). In this work, we report on a crucial first step in applying such methods to discourse samples. Specifically, we describe the results of a computational model that analyzes the context in which a paraphasia occurs in a discourse sample and predicts the speaker’s intended word (or a set of possible intended words). Below, we describe the key role that this specific task of target word prediction plays in the clinical assessment of discourse samples from PWA, motivate our overall computational approach, and describe our model and its behavior. In addition, we evaluate the impact of clinical features of the speaker on our model’s ability to correctly predict target words. This part of the work highlights specific areas where current technology falls short and points to missing pieces that the field must address.

## Assessing Anomia at Discourse Level

It is well documented in the literature that the ability to produce discourse is what matters most to PWA and their families (Cruice et al., 2003; Mayer & Murray, 2003). Yet, despite their popularity, there is evidence that confrontation naming tests cannot fully account for the severity and patterns of anomia exhibited during connected speech. First, connectionist accounts of word retrieval at the discourse level highlight how lexical characteristics of target words interact with activated representations within and across different linguistic levels (e.g., phonological, semantic) (Bock, 1995; Dell, 1986; Dell et al., 1999; Schwartz et al., 2006; Levelt, 1999; Levelt et al., 1999). In addition, several models (e.g. MacDonald, 1994; Tabor et al., 1997) emphasize the influence and relative strength of naturally occurring probabilistic constraints in language use on the activation of linguistic representations. In fact, there seems to be a general consensus in recent empirical investigations that while performance in confrontation naming tests is related to discourse-level performance, analyzing discourse directly may provide unique and useful clinical insights not gained via confrontation naming tests (Fergadiotis et al., 2019; Hickin et al., 2001; Mayer & Murray, 2003; Pashek & Tompkins, 2002). Therefore, relevant assessment tools for aphasia should a) operate at the discourse level, b) be able to capture changes in language skills over time, and c) be routinely included as therapy outcome measures.

At the level of single words, anomia severity is commonly assessed using picture naming tests and reported in terms of overall accuracy scores or ability estimates. Further, a more in-depth analysis of the types and frequencies of word production errors can reveal which linguistic processes that support word access and retrieval are more or less disrupted (Dell et al., 1997). Theoretical accounts of word production allow professionals and/or algorithms to classify an individual’s collection of paraphasias in order to create a detailed profile of that individual’s anomia. This paraphasia classification process requires a series of binary judgments with regards to the paraphasia and its relationship to the intended target word. Specifically, those judgments are: 1) lexicality, i.e., whether or not the paraphasia is a real word; 2) semantic similarity, i.e., whether or not the paraphasia is semantically related to the target; and 3) phonological similarity, i.e., whether or not the paraphasia is phonologically related to the target. To highlight a couple of classification examples, a Semantic paraphasia is a real word that is semantically related to its intended target but phonologically unrelated (e.g., “beard” for “mustache”); whereas a neologism is a nonword, not semantically related by definition, that is phonologically related to the target (e.g., “mustaff” for “mustache”). Lexical or real word paraphasias are understood to represent mostly impairments in lexical-semantic access while nonword paraphasias are thought to reflect deficits in phonological encoding. To help make this time– and labor-intensive assessment process more efficient and therefore more feasible for clinical settings, our research team has developed a paraphasia classification algorithm called ParAlg (Paraphasia Algorithms) that automatically classifies word production errors in the context of object picture naming tests (Casilio et al., 2023; Salem et al., 2022; Fergadiotis et al., 2016; McKinney-Bock & Bedrick, 2019). ParAlg’s paraphasia classifiers algorithmically mirror the main paraphasia classification criteria of the Philadelphia Naming Test (Roach et al., 1996), which includes one of the most well-established and thorough frameworks for error classification during object picture naming.

The accuracy of this multistep paraphasia classification process, however, is entirely predicated on successfully identifying a given paraphasia’s intended target. Target identification is relatively straightforward in the context of confrontation picture naming tests, where the target is presumed to be the word depicted in the picture, but in the context of discourse, determining the target is not as straightforward. Researchers and clinicians undertake this task by applying background knowledge of word production disorders and common anomic patterns (Martin, 2017), as well as general knowledge of the discourse task itself, such as the expected lexicon and the expected temporal arrangement of that lexicon given the overall narrative structure. Furthermore, target prediction can incorporate a multitude of localized contextual factors such as timely gestures, re-tracings from the paraphasia to or toward the intended target, phonological fragments or false starts leading up to the paraphasia, syntactic/semantic information immediately surrounding the paraphasia, and/or semantic and phonological similarities between the paraphasia and its working hypothesis target.

In light of this highly variable and complex process, the preliminary focus of this automation work and of the current paper is to leverage and model the semantic information surrounding word production breakdowns. Elegantly enough, this approach mirrors widely accepted models of spoken word production, such as Dell’s model described earlier where step one involves identification and activation of semantic representations surrounding the target word. One additional and imminent aim of this work, though outside of the scope of this paper, is the exploration of a more fully-automated and naturalistic application of ParAlg – classification of paraphasias in discourse using machine-generated targets. While the present paper explores automatic target prediction for a full range of content words (nouns, verbs, adverbs, adjectives), we do not anticipate being able to classify paraphasias with non-noun targets until equally robust psycholinguistic models are developed for additional parts of speech.

## Novel Approaches for Assessing Paraphasias at Discourse Level

Given the resource-intensive nature of discourse analysis, several computational approaches have been developed to assist researchers and clinicians in analyzing discourse such as automated speech and language measures (e.g., Fergadiotis & Wright, 2011; Bryant et al., 2013; Miller & Iglesias, 2012; Forbes et al., 2014; Day et al., 2021; Chatzoudis et al., 2022). An active area of research is establishing automatic speech recognition (ASR) systems that are effective on aphasic speech (e.g., Le & Provost, 2016; Perez et al., 2020; Gale et al., 2022), some of which are developed and used for diagnosing aphasia or aphasia subtypes (e.g., Fraser et al., 2013; Le et al., 2018). Some preliminary attempts have been made at automated classification of paraphasias in connected speech, but these studies have focused solely on the task of *detecting* paraphasias and determining if they are real words or neologisms (Le et al., 2017; Pai et al., 2020), as opposed to complete classification. Despite the recent advances in automated approaches, to this date there are no computer assisted discourse analyses for the identification and fine-grained classification of paraphasias, the production of which is the hallmark characteristic of most PWA.

Our first attempts at predicting targets of paraphasias in discourse were made using more traditional n-gram and early neural net based language models (Adams et al., 2017), but since then, there have been significant developments in the field of language modeling. In this work, to automatically predict the intended targets of paraphasias in discourse using the surrounding language, we use a machine learning-based transformer language model. Transformer models were first introduced in 2017 (Vaswani et al., 2017) and have since become ubiquitous in NLP research due to their high performance; their structure allows them to be trained on large scale datasets with graphical processing units (GPUs). The introduction of transformer models led to the development of BERT (Bidirectional Encoder Representations from Transformers; Devlin et al., 2019), a large language model (LLM) which has been successful on a variety of NLP tasks such as Google search, text summarization, and question answering (Devlin et al., 2019; Liu & Lapata, 2019; B. Schwartz, 2020). BERT is designed to be pre-trained on a very large scale general purpose dataset and can then be used in its out-of-the-box pre-trained format, or one can use transfer learning to adapt them for a specific domain and task with a process called fine-tuning. During fine-tuning, the model is trained further on a downstream task with domain-specific data. This process allows the models to work well even on tasks with fewer data resources (Zaheer et al, 2021).

LLMs have been successfully applied to a variety of biomedical language tasks. For example, by fine-tuning BERT with PubMed abstracts and clinical notes, Peng et al. (2019) outperformed previous state-of-the-art on five biomedical tasks (e.g., similarity of two sentences from Mayo Clinic clinical data). Researchers have also found success applying these models to clinical language research. For instance, Balagopalan et al. (2020) fine-tuned BERT to detect Alzheimer’s disease from transcribed spontaneous speech. They found that BERT performed better than a standard model based on hand-crafted features. Gale et al. (2021) fine-tuned a variation of BERT called DistilBERT (Sanh et al., 2019) to automatically score commonly used expressive language tasks on a diverse group of children (Autism Spectrum Disorder, Attention-Deficit Hyperactivity Disorder, Developmental Language Disorder, and typical development; age 5-9 years) with high accuracy (83-99%). In previous work developing ParAlg, our group fine-tuned DistilBERT to automatically determine the semantic similarity of lexical paraphasias to the target word with 95.3% accuracy (Salem et al., 2022).

While models like BERT have been very successful, one drawback is that they are designed for relatively short sequences of words; in fact, BERT has a hard limit of taking sequences of text of maximum length 512 tokens. Our data, which consists of retellings of the Cinderella story, includes many sessions longer than that limit. In this work, we instead use a recent LLM called BigBird (Zaheer et al., 2021) which was specifically designed to address this limitation of BERT. Importantly, BigBird, like its predecessor BERT, was trained using “masked language modeling”, a type of sentence cloze task. In this task, randomly selected words from the corpus are masked (i.e., removed and replaced with a special blank token [MASK]), and the model learns to fill in the blank and predict those masked words using the surrounding context, allowing it to learn what words occur in what contexts. This task is in fact similar to our task at hand: we want to predict what target word a person with aphasia was intending to say, given the context of their discourse. Thus, considering the wide success of LLMs, the adaptation of this model to long sequences, and the similarity of its training process to our task, we hypothesized that BigBird would be a good fit for automatically predicting paraphasia targets in discourse.

Given that the current study represents a novel application of a LLM to data from a clinical population, it is worthwhile to explore factors that might influence the accuracy of that approach. It is generally accepted that PWA represent a heterogeneous group in terms of the nature and severity of deficits exhibited during discourse production. For example, some individuals on the mild end of the ability continuum may present with well-constructed utterances during connected speech with only occasional hesitations and single word paraphasias. On the other hand, people on the more severe end of the distribution may exhibit morphosyntactic disturbances as well as significant manifestations of word retrieval deficits including abandoned phrases, revisions, retracings, reformulations, as well as multiple paraphasias. Therefore, given that the LLM relies on the surrounding context of a masked word for prediction, it is conceivable that the success of the model may depend on overall aphasia severity of the speaker. In addition to overall aphasia severity, the predictive utility of the LLM may also depend on the nature of the syntactic deficits exhibited by people with aphasia. Specifically, connected speech from PWA can be characterized as agrammatic or paragrammatic (Butterworth & Howard, 1987; Goodglass, 1993; Saffran et al., 1989; Thompson et al., 1997). Agrammatic speech is typically characterized by an overall reduction of grammatical morphology, simplification of syntactic structure, and overreliance on content words, primarily nouns. On the other hand, paragrammatism is associated with misuse of grammatical aspects including inflectional morphology, significant word substitutions that cross word class, as well as pronounced errors in word ordering. Finally, during discourse production, there are instances where a speaker’s intended target is clear, but that is not always the case, and different raters can disagree. In this study, in addition to clinical factors, we investigated the performance of our LLM as a function of the certainty with which raters can perform the same task.

## Purpose of Study

The purpose of the current study was to create a baseline model for automated target word prediction of paraphasias within spoken discourse using the surrounding language alone. We fine-tuned the LLM BigBird to predict the intended target word of paraphasias within transcripts of the Cinderella story retell task using data from controls, PWA, and a combination. We compared the various models’ accuracy at predicting the correct target word that the human raters identified. We hypothesized that fine-tuning the LLM using task data from control participants as well as PWA would lead to the highest accuracy. Additionally, we evaluated the impact of clinical characteristics and human certainty of target prediction on the model performance. These aims can be summarized in two research objectives: 1) assess the feasibility of applying a modern LLM to this task and establish a performance baseline; 2) explore the impact of clinical factors (specifically fluency and aphasia severity) and intended target ambiguity (according to human raters) on model performance.

## Method

### Data

Data consisted of 353 Cinderella story retelling transcripts from 254 PWA from the English AphasiaBank database (MacWhinney et al., 2011). In this task, participants are first given a wordless picture book of the Cinderella fairytale to briefly review, and then are given a few minutes to recite the story from memory. Demographic and clinical information on these 254 participants at their first session is shown in Table 1. We also supplemented this data with 256 transcripts from control participants without aphasia in AphasiaBank. Our data preparation pipeline is illustrated in Figure 1. More details are provided in the sections below.

**Figure 1.**
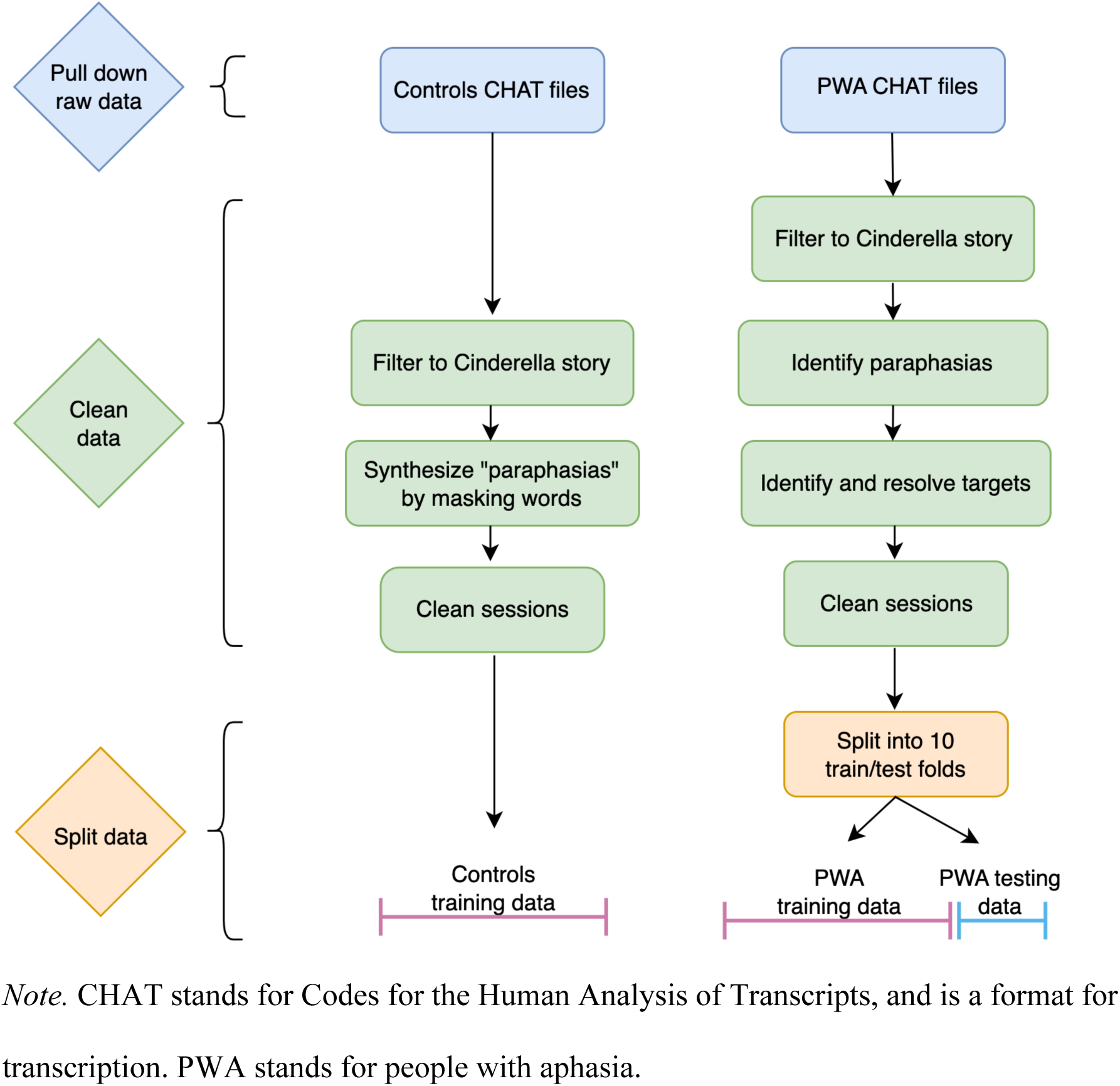
Data preparation pipeline.

**Figure 2.**
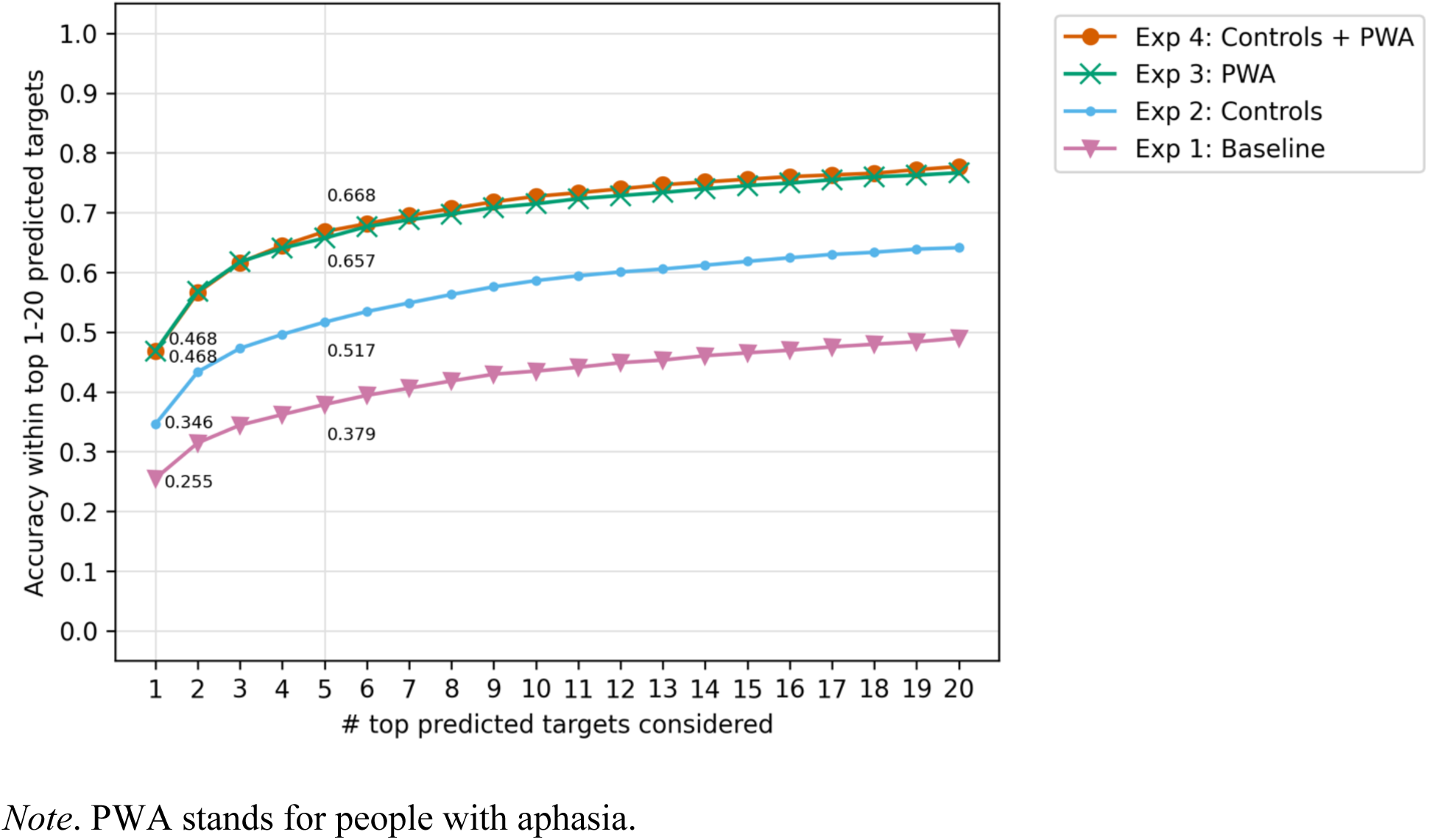
Accuracy within top 1-20 predicted targets for experiments 1-4.

**Table 1.**
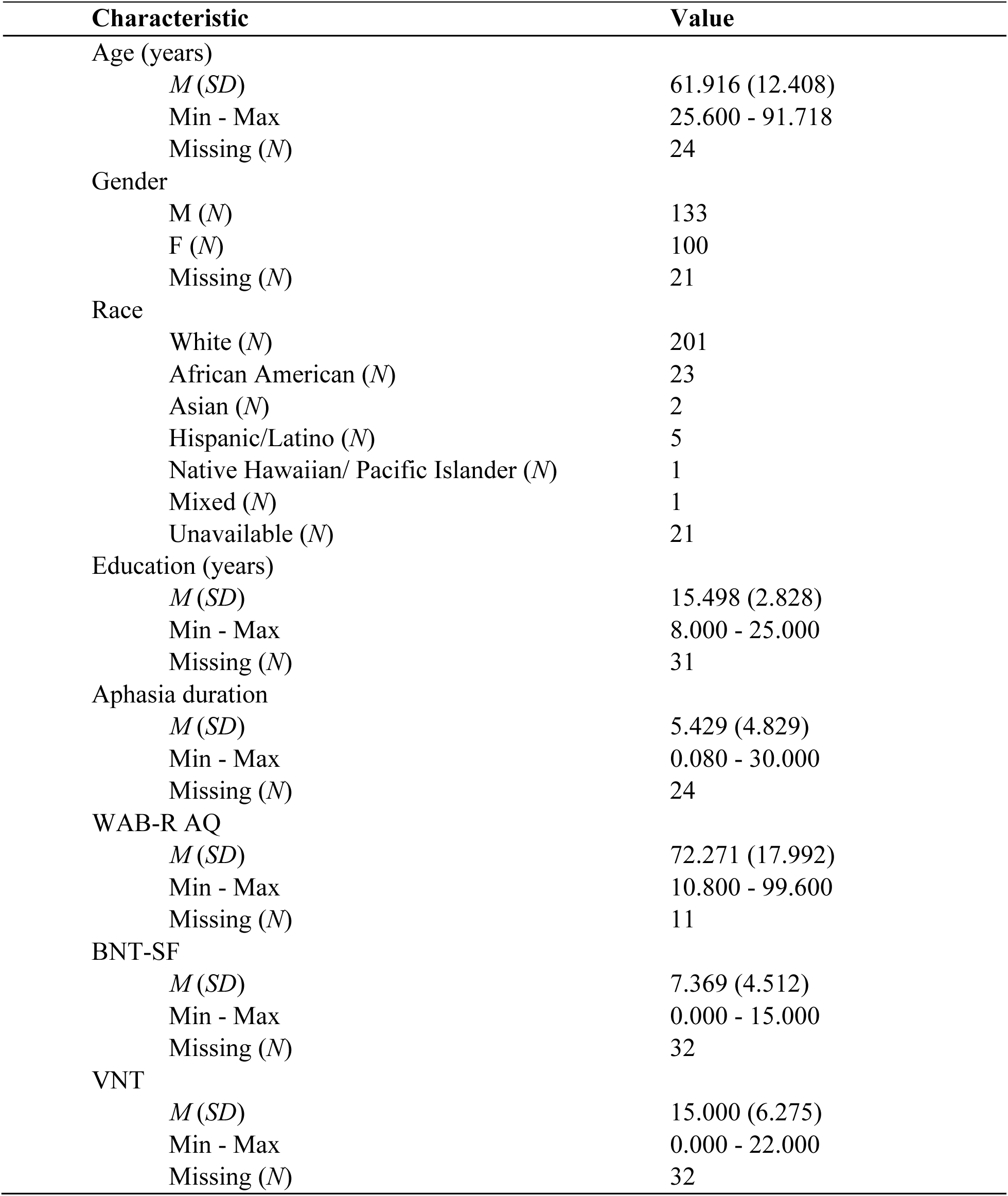
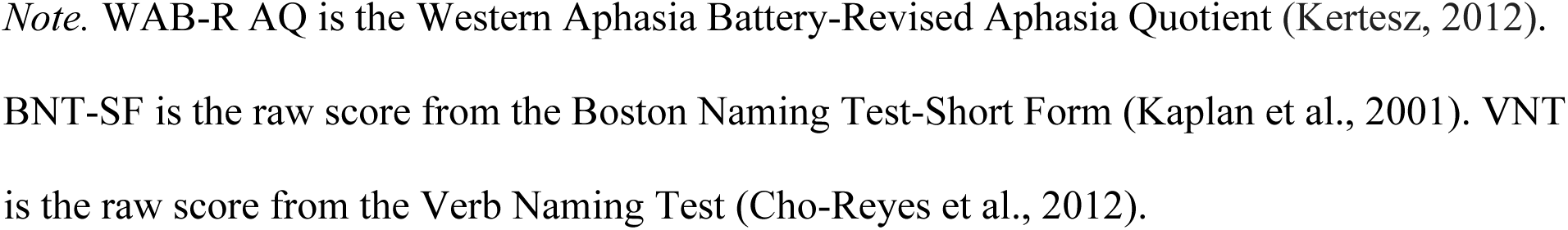
Clinical and demographic information for the 254 participants at their first session.

| Characteristic | Value |
| --- | --- |
| Age (years) |  |
| *M (SD)* | 61.916 (12.408) |
| Min - Max | 25.600 - 91.718 |
| Missing (*N*) | 24 |
| Gender |  |
| M (*N*) | 133 |
| F (*N*) | 100 |
| Missing (*N*) | 21 |
| Race |  |
| White (*N*) | 201 |
| African American (*N*) | 23 |
| Asian (*N*) | 2 |
| Hispanic/Latino (*N*) | 5 |
| Native Hawaiian/ Pacific Islander (*N*) | 1 |
| Mixed (*N*) | 1 |
| Unavailable (*N*) | 21 |
| Education (years) |  |
| *M (SD)* | 15.498 (2.828) |
| Min - Max | 8.000 - 25.000 |
| Missing (*N*) | 31 |
| Aphasia duration |  |
| *M (SD)* | 5.429 (4.829) |
| Min - Max | 0.080 - 30.000 |
| Missing (*N*) | 24 |
| WAB-R AQ |  |
| *M (SD)* | 72.271 (17.992) |
| Min - Max | 10.800 - 99.600 |
| Missing (*N*) | 11 |
| BNT-SF |  |
| *M (SD)* | 7.369 (4.512) |
| Min - Max | 0.000 - 15.000 |
| Missing (*N*) | 32 |
| VNT |  |
| *M (SD)* | 15.000 (6.275) |
| Min - Max | 0.000 - 22.000 |
| Missing (*N*) | 32 |

*Note.* WAB-R AQ is the Western Aphasia Battery-Revised Aphasia Quotient (Kertesz, 2012). BNT-SF is the raw score from the Boston Naming Test-Short Form (Kaplan et al., 2001). VNT is the raw score from the Verb Naming Test (Cho-Reyes et al., 2012).

### Paraphasia Identification

Archival audiovisual recordings and CHAT transcript files (Codes for the Human Analysis of Transcripts; MacWhinney, 2000) of the Cinderella story retell task were retrieved from the English AphasiaBank database on May 4, 2022 for any and all PWA whose sample contained at least one word-level error as annotated by AphasiaBank.^1^ We defined paraphasias as word-level errors made to the lemma of content words (i.e., nouns, verbs, adjectives, adverbs) and excluded from target prediction all other kinds of word-level errors, including those related to disfluency, morphological markings (e.g., plurality, tense), and non-content words (e.g., articles, pronouns). Referencing the CHAT manual (MacWhinney, 2000) accessed on April 13, 2022, we developed a list of word-level error codes for preliminary inclusion and exclusion.

### Target Identification

Target words were identified and annotated in ELAN transcription software (version 6.2), using custom generated templates that also allowed for review of the retellings’ transcripts as well as playback of audiovisual recordings. To maximize transcript readability and efficacy for this task, AphasiaBank transcripts were preprocessed to remove from view additional annotations irrelevant to the task (e.g., utterance-level error coding) as well as the original annotator’s target prediction, if provided.

Target word identifications were completed by five trained student research assistants in pseudorandom order under the supervision of a research SLP, resulting in a total of three independent target identifications for each paraphasia. Research assistants were instructed to watch the audiovisual recordings of the Cinderella story retell task and make their paraphasia target predictions based on a number of contextual factors, including background knowledge related to word production disorders and the Cinderella story. For each identified target, a confidence rating ranging from 1 to 4 was assigned with 1 signifying very unconfident, 2 unconfident, 3 confident, and 4 very confident. In the process, research assistants flagged for potential exclusion any word errors believed to be outside the scope of this project (e.g., the predicted target is not a noun, verb, adjective, or adverb) or produced in the context of personal commentary (e.g., a comment about the difficulty of the task, performance on the task, etc.).

Identified targets from our research assistants as well as AphasiaBank annotators were automatically extracted and compiled for side-by-side comparison and resolution in a spreadsheet. Discrepancies in target words and word errors flagged for exclusion were resolved by a research SLP to arrive at a single, best target identification and in some cases multiple viable target words were provided (e.g., shoe vs. slipper, coach vs. carriage). If there was universal agreement among all three raters and AphasiaBank, then that target was not subject to resolution. If there was disagreement among raters, rater confidence was low, and the resolver could not arrive at a suitable prediction upon review, then the target was listed as “unknown”. All paraphasia-target pairs were reviewed by the research SLP for phonological similarity and whether or not an intermediary target was readily apparent (e.g., the paraphasia “bot”, where “bot” could be interpreted as phonemic paraphasia of “boot”, the intermediary target, and “boot” could be interpreted as a semantic paraphasia of “slipper”, the ultimate target). We calculated average confidence scores (between the three research assistants) and percent agreement (between the three research assistants and the original AphasiaBank target, where available) for each identified target. After filtering to content word paraphasias and excluding paraphasias with unknown targets, we were left with 353 Cinderella story sessions from 254 participants, with a total of 2489 paraphasias.

### Session Text Cleaning

We compiled our target identifications as well as human rater confidence and percent agreement in the CHAT file format. We added our annotations within the “comment on main line” markers specified in the CHAT manual, formatted in a structured notation (YAML) which can be parsed in common programming languages such as Python. The following example shows one such transcript, with our additional annotations highlighted in boldface type:

> *PAR: and she rode off with the pɪnts@u [: prince] **[% {target: a, agreement: 1.0, confidence: 3.33}]** [* p:n] . •680333_684666•

To prepare the transcripts for use with our LLM, we automated a process to convert the transcripts to a more natural-looking written English. Motivated by the long-term goal of a fully automated anomia system, we generally aimed to prepare the transcripts to look like those an automatic speech recognition system would produce. Markings indicating prosodic (e.g. pauses) and paralinguistic details (e.g. gestures) were removed. The CHAT format also uses special markers to indicate phenomena peculiar to the spoken modality, such as retracing and repeats.

For situations like these, we omitted the special markers, but retained most of the spoken content, though we discarded extraneous words that could be identified by simple rules (e.g. a list of filler words like “um”).

In the AphasiaBank files, the transcripts are segmented into units called “utterances” or “conversational units.” These units look similar to sentences—they are delimited by periods— but tend to be shorter and more fragmentary, owing to the inherent differences between spoken and written language. Especially as compared to the written text used to pre-train LLMs, the utterance segmentation guidelines laid out by the CHAT manual would not reliably contain a substantial amount of semantic context for our masked word prediction task. So, while popular LLMs (e.g. BERT) typically process a sentence or two at a time, our transcripts do not divide cleanly into sentences. Rather than attempt to redraw the AphasiaBank-provided utterance boundaries to suit our task, we chose to prepare our data with a full context. In other words, for each paraphasia shown to the LLM, the model was working with a participant’s complete retelling of the Cinderella story.

Each paraphasia was prepared for training or testing by replacing it with a “blank” token (also known as a “mask”) and filling in the other paraphasias in the session with the human identified target word. The following example from above illustrates the cleaned sentence in context, where the paraphasia has been replaced with a mask token:

> … and then and and she put her foot in the. and she rode off with the **[MASK].** Cinderella was pretty girl. …

During fine-tuning and testing, the model learned to fill in the blank of the mask token with the most likely word given the context of the rest of the Cinderella story retelling.

### Data Splitting

We used ten-fold cross validation of the PWA data in order to reduce model overfitting. That is, we divided the 2,489 instances into ten groups and trained ten separate models for each experiment, in each of which one group was held out as testing data. This was done in such a way that for each of the ten iterations, a participant’s responses were only in either the training data or the testing data to prevent the models from learning participant-specific information, and the distribution of Western Aphasia Battery-Revised (WAB-R; Kertesz, 2007) Aphasia Quotient (AQ) scores in training and testing was as close as possible. When evaluating overall performance, the results from the ten test set splits were concatenated, and performance on the entire set of 2489 paraphasias was examined. The same ten-fold splits were used for all experiments.

### Control Data Augmentation

To add additional training data for our experiments and reduce overfitting, we conducted data augmentation (a method of adding synthetic data; see Feng et al., 2021 for more background) on sessions of the Cinderella retelling task from control participants without aphasia. We retrieved all files in AphasiaBank from control participants with a Cinderella story task on April 12, 2022 and added synthetic paraphasias to these sessions. For each session, for each utterance spoken by the participant, with a 20% chance we randomly assigned a content word (one of: noun, verb, adjective, adverb) to be a “paraphasia” to be predicted. This left a control dataset with 256 sessions from 248 participants, with a total of 2427 synthetic paraphasias, which was very close to the number of paraphasias from the PWA data (2489). We cleaned and prepared these sessions using the same process as for PWA data, described in the subsection Session Text Cleaning.

## Model Training and Experiments

In all experiments we used a pre-trained version of the LLM BigBird (Zaheer et al., 2021). This model is a machine learning-based transformer model. Specifically, it is a sparse-attention version of BERT designed for longer sequences of text. As previously mentioned, it was pre-trained on masked language modeling. During masked language model training, the model is given sentences from the corpus where 15% of the tokens are masked (i.e., removed and replaced with a special non-word token, “[MASK]”), and the model attempts to predict what those masked words were given the context of the surrounding sentence. By doing this on the whole corpus of sentences, the model learns what words occur in what contexts. We accessed this pre-trained BigBird from the HuggingFace transformer library (Wolf et al., 2020).

For each experiment (excluding the baseline experiment), we fine-tuned the LLM using another masked language modeling task. Specifically, given the context of the whole Cinderella story transcript, the model tried to fill in the blank of the mask token with the intended target.^2^ The model then compared that prediction with the human-determined ground truth intended target (or the original word for control participants), and learned from its correct and incorrect predictions. The fine-tuning process was repeated on the whole training data set until early-stopping occurred, meaning performance stopped improving on a small portion of the testing data that was held out. Once the model was fine-tuned, we tested it on the PWA paraphasias, which were prepared in the same way as the training data, with each paraphasia sequentially replaced with a mask, and all others filled in with their target. At test time, we pulled out the model’s top prediction, as well as its nineteen next most likely predictions, giving us its top twenty predictions for the target, sorted from most likely to least likely. We considered more than just the top prediction because there is inherent ambiguity in target identification, and in future work we may consider multiple possible targets when classifying paraphasias in discourse.

We conducted four experiments using different preparations of training data, which are summarized in Table 2. In Experiment 1, we used the pre-trained BigBird model without any fine-tuning using Cinderella story data. We considered this our “baseline” model to beat. In Experiment 2, we fine-tuned the LLM using just the Cinderella story sessions from control participants with synthetic paraphasias. In Experiment 3, the pre-trained model was fine-tuned using Cinderella story sessions from PWA. Finally, in Experiment 4, the model was fine-tuned using a combined data set of control participant data *and* PWA data.

**Table 2.**
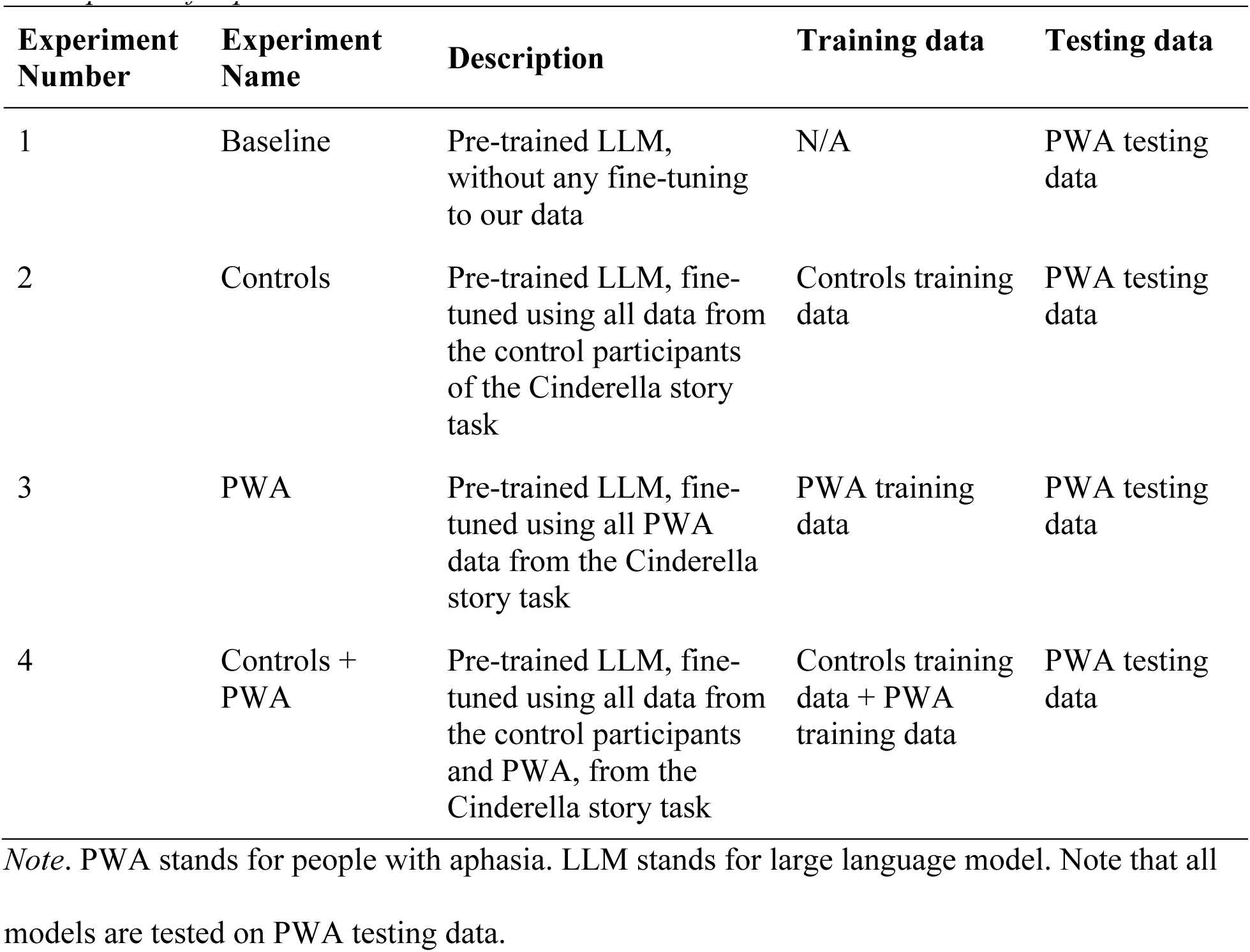
Descriptions of experiments 1-4.

| Experiment Number | Experiment Name | Description | Training data | Testing data |
| --- | --- | --- | --- | --- |
| 1 | Baseline | Pre-trained LLM, without any fine-tuning to our data | N/A | PWA testing data |
| 2 | Controls | Pre-trained LLM, fine-tuned using all data from the control participants of the Cinderella story task | Controls training data | PWA testing data |
| 3 | PWA | Pre-trained LLM, fine-tuned using all PWA data from the Cinderella story task | PWA training data | PWA testing data |
| 4 | Controls + PWA | Pre-trained LLM, fine-tuned using all data from the control participants and PWA, from the Cinderella story task | Controls training data + PWA training data | PWA testing data |

*Note.* PWA stands for people with aphasia. LLM stands for large language model. Note that all
models are tested on PWA testing data.

## Evaluation

We evaluated performance of the four experiments using accuracy. We calculated the accuracy of “exact match” between the model’s top predicted intended word and the human determined target word by counting up the number of matches and dividing by the total number of test instances. Additionally, we calculated the accuracy within the top one-20 model predictions. That is, we counted up how many times out of all test instances the human determined target word was: the top model prediction (i.e., top one or exact match); the first or second model prediction (top two); the first, second or third model prediction (top three); and so on for up to 20 chances to predict the right target. We primarily compared accuracy within one chance (exact match) and accuracy within five chances for the four experiments. We determined whether disagreements between exact match accuracy of the models were significant using McNemar’s test with continuity correction (McNemar, 1947).

First, we calculated accuracy on all 2489 paraphasias. To determine what factors influenced model performance, we also calculated exact match and within five accuracy on several different test set stratifications for each model. We calculated performance separately on sessions from participants with WAB-R AQ above or below the median, participants with fluent aphasia (Wernicke, Anomic, Conduction, or Transcortical Sensory aphasia, or those considered “non aphasic” by the WAB-R) and non-fluent aphasia (Broca, Global, or Transcortical Motor aphasia), test instances where the human raters had high confidence (above median) or low confidence (below median) in intended target determination, and test instances where human raters had perfect agreement in determining the intended target, or imperfect agreement. We tested whether differences in performance between these stratifications were significant using two-sided z-tests for independent proportions. Throughout, a *p*-value of <0.05 was retained as a level of statistical significance.

## Results

Accuracy results from Experiments 1-4 are shown in Tables 3, 4, 5, and 6, respectively. Experiment 1, our baseline model, achieved 25.5% for exact match accuracy on all paraphasias. Experiment 2, the model fine-tuned on control data, achieved 34.6% exact match accuracy. Experiments 3 and 4 (fine-tuned on PWA data and controls plus PWA data respectively) both achieved exact match accuracy of 46.8%, 21.3 points above the baseline model. According to McNemar’s test, Experiment 3 and Experiment 4’s exact match accuracy levels were significantly different than both Experiment 1 (the baseline model) and Experiment 2, all with *p* < 0.001. Experiment 3’s exact match accuracy was not significantly different from Experiment 4’s exact match accuracy (*p* = 0.963).

**Table 3.** Experiment 1: Baseline.

| Test set | Number of paraphasias | Accuracy exact match | Accuracy within 5 |
| --- | --- | --- | --- |
| All paraphasias | 2489 | 0.255 | 0.379 |
| Human agreement = 100% | 1244 | 0.309 | 0.405 |
| Human agreement < 100% | 1245 | 0.201 | 0.353 |
| Human confidence > median (3.3) | 1089 | 0.319 | 0.419 |
| Humans confidence <= median (3.3) | 1400 | 0.206 | 0.348 |
| WAB-R AQ > median (74.6) | 1039 | 0.294 | 0.410 |
| WAB-R AQ <= median (74.6) | 1076 | 0.204 | 0.325 |
| Fluent participants | 1666 | 0.261 | 0.385 |
| Non-fluent participants | 449 | 0.198 | 0.301 |

*Note.* WAB-R AQ is the Western Aphasia Battery-Revised Aphasia Quotient (Kertesz, 2012).
Fluent participants are those with Wernicke, Anomic, Conduction, or Transcortical Sensory
aphasia, or those considered “non aphasic” by the WAB-R. Non-fluent participants are those
with the Broca, Global, or Transcortical Motor aphasia. 48 out of 353 total sessions had
unavailable WAB-R results and were excluded just from analyses involving WAB-R scores.
Accuracy exact match refers to the top model prediction of target word matching the human-
identified target word. Accuracy within 5 refers to the human-identified target word being one of
the top five model predictions.

**Table 4.** Experiment 2: Fine-tuned on controls data.

| **Test set** | **Number of paraphasias** | **Accuracy exact match** | **Accuracy within 5** |
| --- | --- | --- | --- |
| All paraphasias | 2489 | 0.346 | 0.517 |
| Human agreement = 100% | 1244 | 0.436 | 0.600 |
| Human agreement < 100% | 1245 | 0.255 | 0.434 |
| Human confidence > median (3.3) | 1089 | 0.453 | 0.614 |
| Humans confidence <= median (3.3) | 1400 | 0.263 | 0.441 |
| WAB-R AQ > median (74.6) | 1039 | 0.398 | 0.580 |
| WAB-R AQ <= median (74.6) | 1076 | 0.290 | 0.453 |
| Fluent participants | 1666 | 0.362 | 0.543 |
| Non-fluent participants | 449 | 0.274 | 0.414 |

*Note.* WAB-R AQ is the Western Aphasia Battery-Revised Aphasia Quotient (Kertesz, 2012).
Fluent participants are those with Wernicke, Anomic, Conduction, or Transcortical Sensory aphasia, or those considered “non aphasic” by the WAB-R. Non-fluent participants are those with the Broca, Global, or Transcortical Motor aphasia. 48 out of 353 total sessions had unavailable WAB-R results and were excluded just from analyses involving WAB-R scores. Accuracy exact match refers to the top model prediction of target word matching the human-identified target word. Accuracy within 5 refers to the human-identified target word being one of the top five model predictions.

**Table 5.** Experiment 3: Fine-tuned on PWA data.

| Test set | Number of paraphasias | Accuracy exact match | Accuracy within 5 |
| --- | --- | --- | --- |
| All paraphasias | 2489 | 0.468 | 0.657 |
| Human agreement = 100% | 1244 | 0.595 | 0.767 |
| Human agreement < 100% | 1245 | 0.342 | 0.548 |
| Human confidence > median (3.3) | 1089 | 0.605 | 0.768 |
| Humans confidence <= median (3.3) | 1400 | 0.362 | 0.571 |
| WAB-R AQ > median (74.6) | 1039 | 0.527 | 0.703 |
| WAB-R AQ <= median (74.6) | 1076 | 0.416 | 0.621 |
| Fluent participants | 1666 | 0.487 | 0.670 |
| Non-fluent participants | 449 | 0.412 | 0.626 |

*Note.* PWA stands for people with aphasia. WAB-R AQ is the Western Aphasia Battery-Revised Aphasia Quotient (Kertesz, 2012). Fluent participants are those with Wernicke, Anomic, Conduction, or Transcortical Sensory aphasia, or those considered “non aphasic” by the WAB-R. Non-fluent participants are those with the Broca, Global, or Transcortical Motor aphasia. 48 out of 353 total sessions had unavailable WAB-R results and were excluded just from analyses involving WAB-R scores. Accuracy exact match refers to the top model prediction of target word matching the human-identified target word. Accuracy within 5 refers to the human-identified target word being one of the top five model predictions.

**Table 6.**
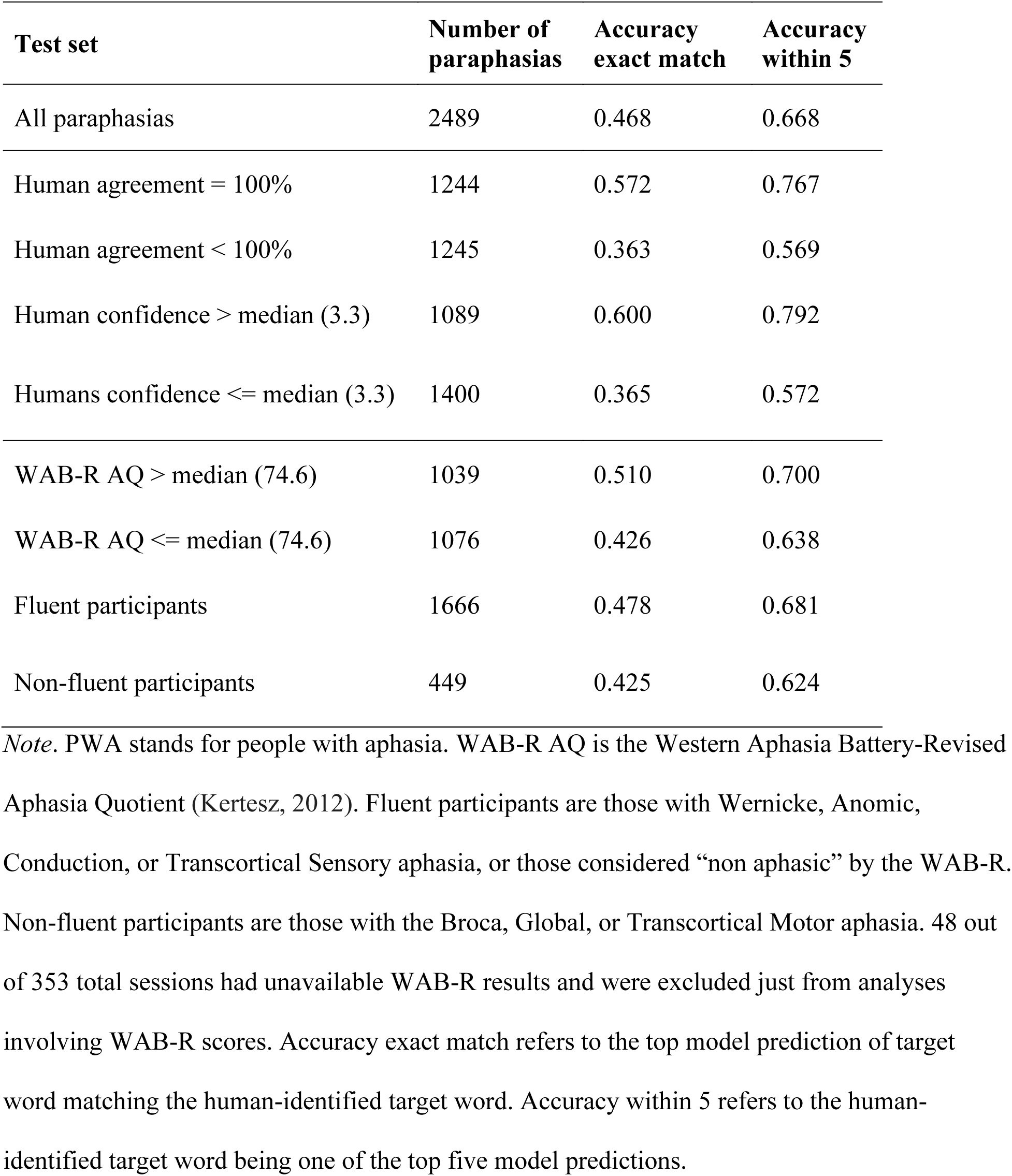
Experiment 4: Fine-tuned on controls and PWA data.

Figure 3 shows accuracy within the top 20 model predictions for all four experiments. Accuracy of all experiments saw the sharpest increase within the top one (exact match) and top five model predictions, and then slower increase when allowing the remaining 15 chances to find the correct target. As stated previously, Experiments 3 and 4 achieved the highest performance of 46.8% exact match accuracy on all paraphasias. Considering within five accuracy, experiment 4 obtained 66.8% accuracy within its top five predictions, which was just one point higher than Experiment 3, which obtained 65.7% accuracy within top five predictions. Regardless of the number of top predicted targets we considered, the baseline performed the lowest, followed by Experiment 2 (trained on controls), and then the two experiments fine-tuned with PWA data were our highest performing models. When looking across accuracy within top one through 20 predictions, the difference in performance between Experiment 4 (fine-tuned on PWA and controls data) and Experiment 3 (fine-tuned on PWA data) was an increase of just one point or less. These findings indicate that performance between these two models was not significantly different. So, without loss of generality, we discuss Experiment 4 in more detail below.

We explored the impact of clinical factors and intended target ambiguity on model performance by sequentially calculating accuracy of the test set stratified by these factors. Considering exact match accuracy, performance in Experiment 4 was higher (59.5%) on the paraphasias with targets humans all agreed upon and lower (34.2%) on the paraphasias with less than perfect agreement. A similar pattern emerged for human confidence, with higher accuracy (60.5%) on paraphasias with targets humans were more confident at identifying and lower accuracy (36.2%) on targets with lower human confidence. We also saw higher performance on sessions where the participant had a WAB-R AQ higher than the median (52.7% accuracy) versus those where the participant had a WAB-R AQ below the median (41.6% accuracy). Similarly, we saw higher performance on the participants with fluent aphasia (48.7% accuracy) than the participants with non-fluent aphasia (41.2% accuracy). Overall, the highest accuracy out of all test sets was on the paraphasias with high human confidence in target determination. For each of these four comparisons, the two test set stratifications (e.g., perfect human agreement vs imperfect human agreement) obtained significantly different performance levels according to the two-sided *z*-test for independent proportions (see Supplemental Table 1 in the Supplemental Material). *P*-values were all <= 0.001 except for the fluent versus non-fluent stratification, which had *p* = 0.016. The same directions of performance difference were seen for the accuracy within the top five predictions of these comparisons. The highest within-five accuracy out of all test set stratifications was also seen for the above median human confidence paraphasias, which Experiment 4 got correct 76.8% of the time within the top five model predictions.

## Discussion

In this study, we trained a LLM to automatically predict the intended targets for paraphasias in discourse during the Cinderella story retelling task. We tried various training data configurations and our two best performing experiments were fine-tuned using PWA data, with or without controls data, and achieved exact match accuracy 47%, and accuracy within top five predictions between 66-67%. Considering just one of these (Experiment 4, fine-tuned on PWA and controls data), the model performed better on paraphasias which had targets that were easier for humans to identify. It also performed better on paraphasias from participants with less severe aphasia and fluent aphasia. Overall, this work produced a relatively high performing model for automatically determining paraphasia targets in connected speech, while just using the surrounding context.

Our baseline model achieved an overall exact match accuracy of 25.5%. This model, which was not fine-tuned to our data at all, was able to use its general-purpose recognition of language patterns to make some correct predictions, without having been exposed to the specific vocabulary and structure of the Cinderella story retellings. It is likely that the original corpus of text used in pre-training the LLM would have included examples of various forms of the Cinderella story, but to a much lesser degree had it been fine-tuned to it. The model used in Experiment 2, fine-tuned using data from control-group participants with the addition of synthesized paraphasias, improved by almost ten points beyond the baseline model with exact match accuracy 34.6%. In this experiment, the pre-trained LLM was specifically exposed to the vocabulary and structure of the Cinderella story, as well as the general task of filling in words in it, but it was not exposed to any real-world examples of paraphasias. In contrast, Experiment 3, fine-tuned on just PWA data, saw a 21 point increase in exact match accuracy over the baseline model. Thus, training the model for this task required not just exposing the pre-trained model to the vocabulary of the Cinderella story, but also specifically examples of real-world paraphasias that occur in that task. Somewhat surprisingly, the model using both PWA data and controls data (Experiment 4) did not improve beyond the model fine-tuned with just PWA data (Experiment 3). This likely indicates that the PWA data gave enough of that vocabulary knowledge to the LLM, and the controls data did not provide any further information. However, more work could be done to synthesize paraphasias in the controls data to make them more similar to real-world paraphasias. As described in the Control Data Augmentation subsection, we attempted to make them more “realistic” by only making content words paraphasias, but there are other possibilities that could be explored in future work: adding synthetic re-tracings, for example, as well as utilizing psycholinguistic variables (e.g. length in phonemes, frequency of occurrence, imageability, etc.) to produce more realistic synthetic training data.

We found that human certainty about paraphasia targets was associated with model performance. Specifically, our best performing model (Experiment 4) performed significantly better on paraphasias with targets that humans were more confident on or had perfect agreement on. This association is reassuring and acts as a simple validity check, since it indicates that our trained models had an easier time with the more obvious targets. There is inherent ambiguity in determining targets for paraphasias in discourse. Half of the paraphasias had percent agreement below 100%, and in fact, average percent agreement on target identification was 76.8%.

Moreover, this percentage agreement is only on the paraphasias for which we were able to resolve a target and excludes targets where ground truth could not be determined. Considering 76.8% agreement as a stand-in for the obtainable human accuracy on this task, obtaining 46.8% accuracy on paraphasias with known targets appears high. Relatedly, while the LLM was designed to rely exclusively on the surrounding language for its predictions, human raters had access to audiovisual recordings and transcripts and thus were able predict targets utilizing additional sources of information such as phonological similarity and gestures.

We also found that, as expected, Experiment 4 saw significantly different performance between participants with above median severity and below median severity, according to the WAB-R AQ, with exact match accuracy 8.4% higher on participants with less severe aphasia. The exact reason for this difference in performance, whether it be factors such as increased occurrence of abandoned phrasings or multiple paraphasias from more severe participants, could be examined further. Relatedly, Experiment 4 performed significantly better on fluent participants than non-fluent participants. Our fluent (Wernicke, Anomic, Conduction, Transcortical Sensory, or non-aphasic by WAB-R) and non-fluent (Broca, Global, or Transcortical Motor) stratifications acted as a proxy for capturing paragrammatic and agrammatic aphasia types respectively. The non-fluent (and perhaps agrammatic) participants may have harder to identify targets because of a lack of content words and context for the LLM to rely on. However, we recognize limitations with this approach. We had substantially fewer training examples from non-fluent participants (449 paraphasias) than fluent participants (1666 paraphasias), which may have impacted that performance difference. Additionally, classification based on the WAB-R is not perfect as there is both classification error and considerable heterogeneity within groups. Finally, the mapping between fluency types and type of grammatical deficits is not perfect. Nonetheless, these stratifications of the test set provided some clues on what features impact performance and where the models can improve. It is also possible that, particularly with more training data, separate models trained for use on specific types of aphasia could see higher performance and better clinical utility.

After our quantitative analyses, we conducted an informal review of Experiment 4’s output, observing some of the more apparent patterns. Some errors were rather unsurprising, like swapping similar verbs (e.g. “sweeping” for “cleaning”). Others were random and garbled (e.g. “Cinderellaipper” for “slipper”) and obviously a consequence of the text encoding constraints (see Appendix A). Where larger patterns stood out, though, they tended to point to a few peculiarities of the dataset.

For example, about 26% of the samples in our dataset involved paraphasias which AphasiaBank had annotated as part of a “retracing” event. Retracing is when a speaker abandons a segment of speech and then retries that segment again (e.g. “Cinderella <put on> [//] tried on the slipper”). When a target word was involved in a retracing event, our LLM’s top-five accuracy for target prediction increased to 80% (vs. 62% when it was not). Since we fill in all the paraphasia targets except the current target (see Model Training and Experiments) any other paraphasias in the immediate context would have been filled in with the correct target word, which provides an advantage for the task at hand. However, this can also work against the model when a target was not actually a part of a retracing event. Informally, we observed that the model sometimes incorrectly chose a word from the immediate context, predicting a retracing where there was none.

Another peculiarity of our dataset was the storytelling task itself, marked by a Cinderella-centric distribution of target words. Out of the 523 unique target words, about 30% of targets were one of five salient words from the fairy tale (“Cinderella,” “prince,” “slipper,” “ball,” or “godmother”). For the most common word, “Cinderella” (265 examples, 11% of total), the LLM was correct 170 times (64%) within the first guess and 227 times (86%) within five guesses. However, this advantage was largely canceled out when the correct target was not the protagonist’s name: the model incorrectly predicted “Cinderella” 157 times as a first guess, and 443 times as a top-five guess. Looking at a subset of the data unaffected by the above factors, we find 233 samples which had a unique target word (occurring only once) and also were not part of a retracing event. The first-guess accuracy for these samples dropped from 39% to 15% between the baseline and fine-tuned models, respectively.

These three patterns—predicting targets that were repeats from the surrounding context, frequently predicting common words from the task, and having difficulty with more rare words—are all consequences of fine-tuning a model. There is a tradeoff between the desirable outcome of improving performance by following common patterns in the training data and the loss in performance when new data points break that pattern; this is known as the bias-variance tradeoff and is well documented in machine learning literature (Geman et al., 1992; Belkin et al., 2019). We employed techniques to reduce overfitting to the training data (data augmentation, cross validation, early stopping), but more strategies could be explored.

Given the architecture of our LLM, we suspect various utterance-related measures would also influence target prediction accuracy for a given speaker and/or utterance. For example, we would predict that speakers with longer utterances, i.e., mean length of utterance in words, would be supplying the model with more linguistic information and therefore increase the likelihood of target prediction success. Another set of hypotheses relates to the quality of the speaker’s utterances in terms of completeness, percentage of utterances that are complete sentences; correctness, percentage of syntactically and/or semantically correct sentences; complexity, number of embedded clauses per sentence, sentence complexity ratio (Thompson et al., 1995), and verbs per utterance; as well as lexical diversity measures like type-token ratio and vocd (Malvern, Richards, Chipere, & Purán, 2004). As mentioned previously, these factors may further explain why performance was affected by fluency and aphasia severity. All of the aforementioned speaker outcome measures can be automatically calculated using CLAN software (MacWhinney, 2000), and we posit all of them would be positive predictors of target prediction accuracy. To deepen our understanding and interpretation of our results, therefore, a future direction of this work is to employ a generalized linear mixed effects model to test these hypothesizes and quantify the magnitude of any significant predictors.

There are many other future directions for this work. Currently, we achieve 46.8% accuracy at predicting paraphasia targets by just using the text of the story, excluding the paraphasia. However, in many cases the details of the paraphasia itself would provide useful information for determining the target. In future work, we plan to develop a model that uses both the semantic context surrounding the paraphasia as well as the phonemes of the paraphasia itself to further improve predictive utility. Considering the difficulty of the task at hand, our performance using just the surrounding language is surprisingly high. However, as mentioned, the Cinderella retelling task is a highly constrained activity, with a much smaller expected target vocabulary than in standard speech. In the context of test and scale development for clinical assessment, when batteries typically include one or two specific stories, gains due to the constrained nature of the stimuli are advantageous. However, in the future, it could be beneficial to train models for less constrained tasks or more naturalistic speech. Additionally, these findings open up possibilities for novel applications that extend beyond assessment, such as augmentative and alternative communication systems. Finally, as previously mentioned, we intend to eventually extend ParAlg, our automated system for classifying paraphasias, to use it on discourse. This work generates a preliminary model for the first step in that process: automatically identifying the most likely targets for paraphasias in discourse.

## Data Availability

Data is available from AphasiaBank to all members of the AphasiaBank consortium group (https://aphasia.talkbank.org/).

## Acknowledgments

This work was supported by National Institute on Deafness and Other Communication Disorders Grant R01DC015999 (Principal Investigators: Steven Bedrick and Gerasimos Fergadiotis). We would also like to thank Mia Cywinski, Samuel Hedine, Lidiya Khoroshenkikh, Jonathan Madrigal, and Anya Russell for their crucial work identifying paraphasia targets.

## Data Availability Statement

Data from PWA and controls is available from AphasiaBank to all members of the AphasiaBank consortium group (https://aphasia.talkbank.org/).

## Appendix

## Appendix A: Details of Masking and Decoding

To encode our inputs and outputs into a discrete numerical form recognizable to our specific choice of LLM, the text is encoded as sub-word units called SentencePieces (Kudo & Richardson, 2018). For example, the word “slipper” is represented by two tokens: “sl” and “ipper”. The SentencePieces algorithm identifies token boundaries using an unsupervised statistical algorithm, and its outputs reflect patterns of corpus frequency rather than morphology or any other linguistic principle (though, in practice, on English text there is often some incidental overlap with morphology). For most purposes, these SentencePieces and their contents are an implementation detail, encoded and decoded automatically by tools included with the language modeling software. However, the detail is relevant to two of our methodological choices. First, due to input and output constraints imposed by the architecture of the baseline model, each target word was masked with as many [MASK] tokens as corresponded to its SentencePiece-encoded length. Relatedly, upon decoding our model’s target word predictions, the model produced as many SentencePieces as there were [MASK] tokens in the input sequence. In other words, for our present experimental setup, the model could not produce a prediction with too many or too few SentencePieces. Second, for outputs requiring more than one SentencePiece, we decoded the output using a standard technique known as “beam search” (Lowerre, 1976). Given that the number of possible SentencePiece permutations grows exponentially with each additional [MASK] token, a beam search allows us to efficiently identify possible combinations of SentencePieces by estimating conditional probabilities for only the n most likely tokens at each step in the sequence. We used a limit (“beam width”) of n=20 while decoding our model’s output.

## Supplemental Material

**Supplemental Table 1.**
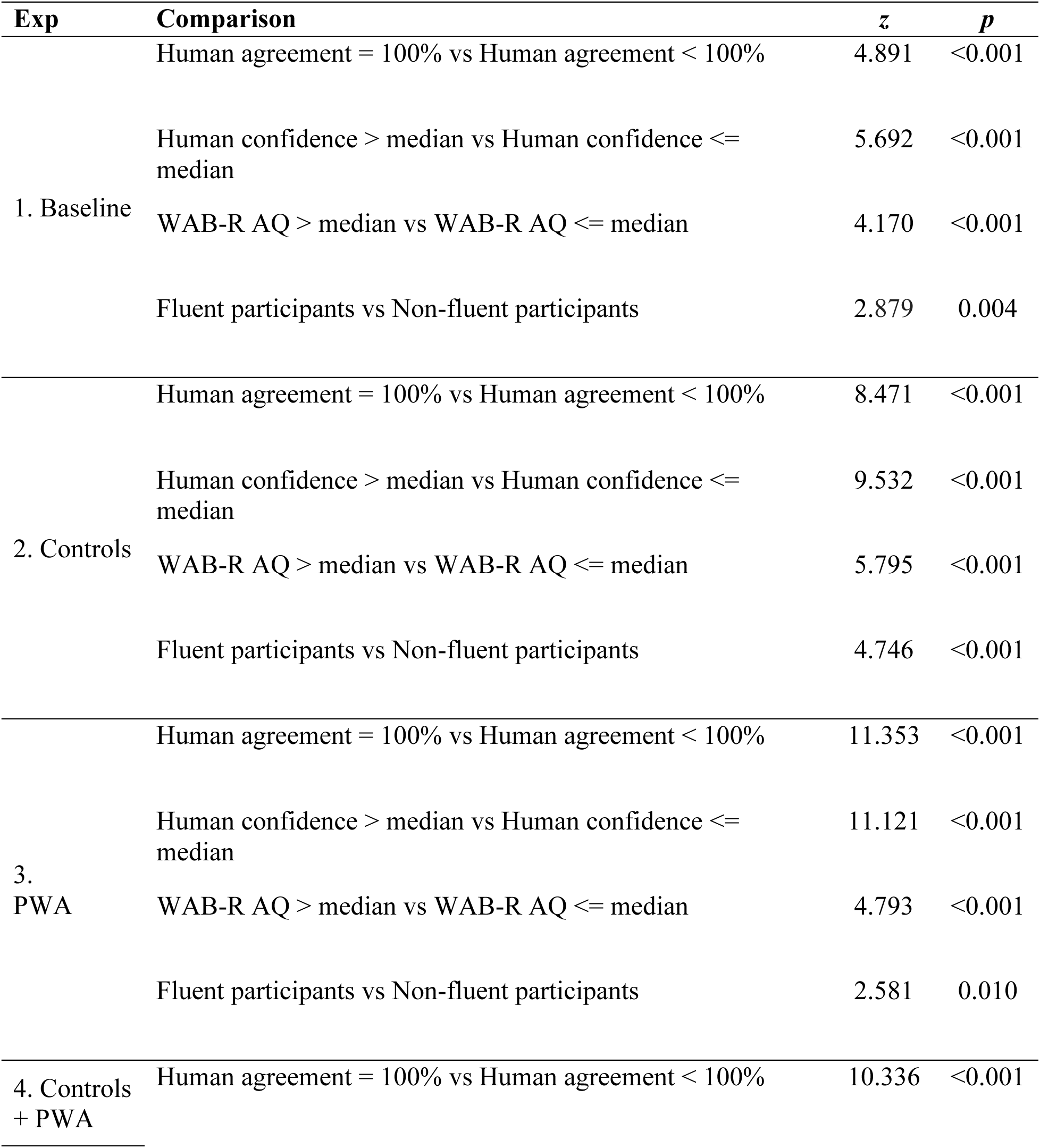
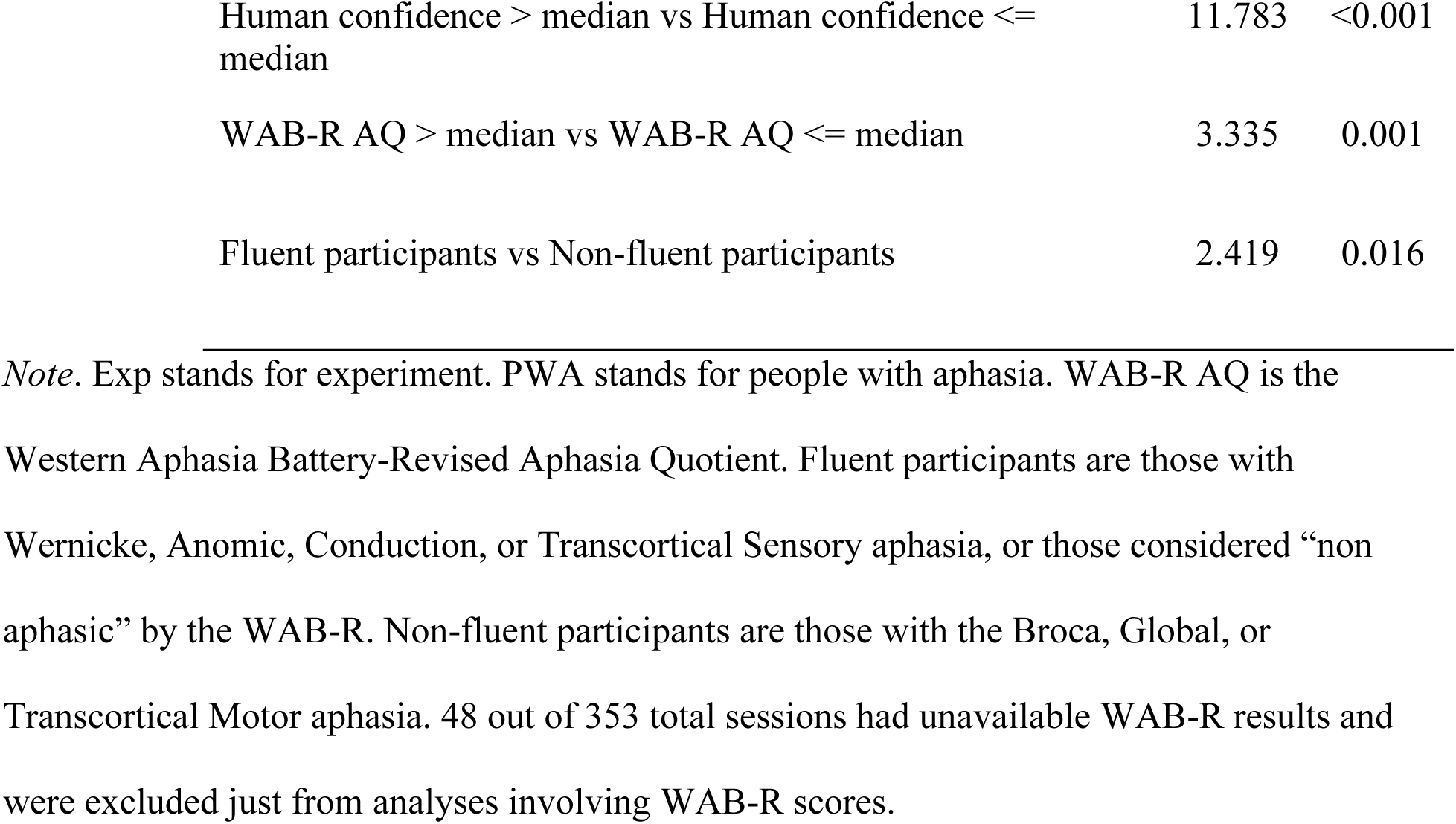
Two-sided z-tests for independent proportions for test set stratifications of exact match accuracy for all experiments.

## Footnotes

1 Although the content of the transcripts is based on the AphasiaBank database on May 4, 2022, we applied updates to the clinical scores that were unavailable on AphasiaBank until December, 2022.

2 There exist certain subtleties to how this is done at a technical level, which we describe in detail in Appendix A. The precise manner in which we performed our masking, and ensuing prediction experiments, would be slightly different had we chosen a different neural model, but the overall methodology would be the same.

